# Perforated appendicitis: can it be a bedside diagnosis?

**DOI:** 10.1101/2020.04.26.20080358

**Authors:** Maham Tariq, Sara Malik, Eesha yaqoob, Mehwish Changez, Saad Javed, Ramlah Ghazanfor, Ghulam Khadija, Javaria Malik, Bilal Ahmad, Khawaja Rafay Ghazanfor

## Abstract

**INTRODUCTION:** Appendicitis remains one of the most common causes of acute abdomen worldwide. It presents as a spectrum of disease ranging from an acutely inflamed appendix to a perforated one. where acutely inflamed can be managed conservatively, a perforated appendix always needs surgery to prevent complications like pelvic abscesses. Bedside diagnosis remains relevant in our setup.

**AIMS AND OBJECTIVES:** To determine whether history, clinical examination, and basic laboratory investigations can help in confident bedside diagnosis of perforated appendicitis especially in the absence of sophisticated diagnostic modalities.

**MATERIALS AND METHODS:** A retrospective case-control study was conducted. Hospital records of patients who underwent open appendectomy in the year 2016 were reviewed. Two groups of 100 patients each were made based on per operative findings. Appendices having macroscopic holes in the base or tip were labeled as perforated. Group A had acutely inflamed appendix and group b had perforated appendix. Patients’ demographic details were taken from hospital admission tickets. Findings of history and examination were retrieved from treating resident and operating surgeon's notes. Data were analyzed through SPSS.

**RESULTS:** Out of 200 patients the total number of males was 102 (51%) and females were 98 (49%). Mean age was 24.13+9.73 in males and 18.7+ 6.4 in females of group A and 26.0+10.1 in males and 20.56+7.53 in females of Group B. Group B showed a significant delay in presentation to emergency after the onset of pain (P = 0.022). Upon history and clinical examination, the presence of anorexia, malaise, generalized abdominal pain, guarding, mass in right iliac fossa were significantly associated with perforation. Whereas gender, fever, vomiting, and dysuria showed no association with perforation.

**CONCLUSION:** Bedside conventional methods of history taking and examination remain a useful tool in anticipating perforated appendicitis. This helps surgeons in planning incisions and prioritizing patients on heavy operating lists. This remains especially relevant in resource-constrained setups where sophisticated modalities like CT scans are largely unavailable.

## INTRODUCTION

Acute appendicitis remains one of the most frequent cases encountered on surgical emergency floors, worldwide. An open or laparoscopic appendectomy was once considered necessary in all cases. Recent guidelines suggest the use of antibiotics in uncomplicated episodes of appendicitis, which is supposed to be equally efficacious and less morbid (1,2). However, the formidable state of perforated and gangrenous appendicitis remains an exception.

Differentiating between a perforated and simple appendicitis happens to be among the commonest parameters studied because of its contribution in guiding treatment and timing of surgery(3) and accuracy in predicting prospective intimidating complications like peritonitis, abscess formation, and postoperative intra-abdominal collection. (4)

Differentiate between a perforated and a non-perforated appendix has been a matter of great debate since both have overlapping presentations. Extremes of ages, increasing duration of symptoms, pyrexia, tenderness outside right lower quadrant, leukocyte count, C-Reactive Protein levels, Erythrocyte Sedimentation Rate levels neutrophil to lymphocyte ratio and high bilirubin count were good predictors of perforation according to several studies. Inturn these parameters will provide a useful guide between the conservative or surgical treatment of appendicitis, and early use of antibiotics. (5,6,7,8,9)

Despite this, a wide range of spectrums of presentation is displayed by both acute and perforated appendix and diagnosing them before surgery remains a dilemma to date. Hence, the importance of meticulous history taking and bedside examination of the patient by the consulting surgeon in clinching the diagnosis cannot be out shadowed. This coupled with lab findings is a useful aid in distinguishing both. Moreover, this can also help in planning incision i.e Gridiron vs Lanz incision, and surgical approach of open vs laparoscopic appendectomy.

Our study was aimed at analyzing the differences in signs, symptoms, and investigations to distinguish an acutely inflamed appendix from a perforated or gangrenous one preoperatively, in essence providing a reliable direction to proceed with.

## MATERIAL AND METHODS

It was a case-control study conducted retrospectively. After permission from the hospital's ethical committee, medical records of all patients who underwent open appendectomy in surgical unit-1, Holy Family Hospital during the year 2016 were retrieved. Patients were separated into two groups: those with perforated and those with non-perforated appendicitis. There were 100 patients in each group. All patients were received in the emergency department by the on-call residents. History taking and clinical examination were performed by the registrars in charge. Per-operative findings documented in operation notes were also recorded. Those cases where a per-operatively macroscopic hole was noted in the appendix were labeled as perforated. Demographic information was collected from hospital admission forms. Data were analyzed by SPSS version 20. Chi-square and independent-sample t-test were used to analyze nominal and categorical data respectively. A p-value of <0.05 was considered statistically significant.

## RESULTS

Two groups of 100 patients each were made. Group A had acutely inflamed appendix and group B had perforated appendix. The total number of males was 102 (51%) and females were 98 (49%). Demographic information of participants is given below in chart no 1.0

**Chart no 1:**
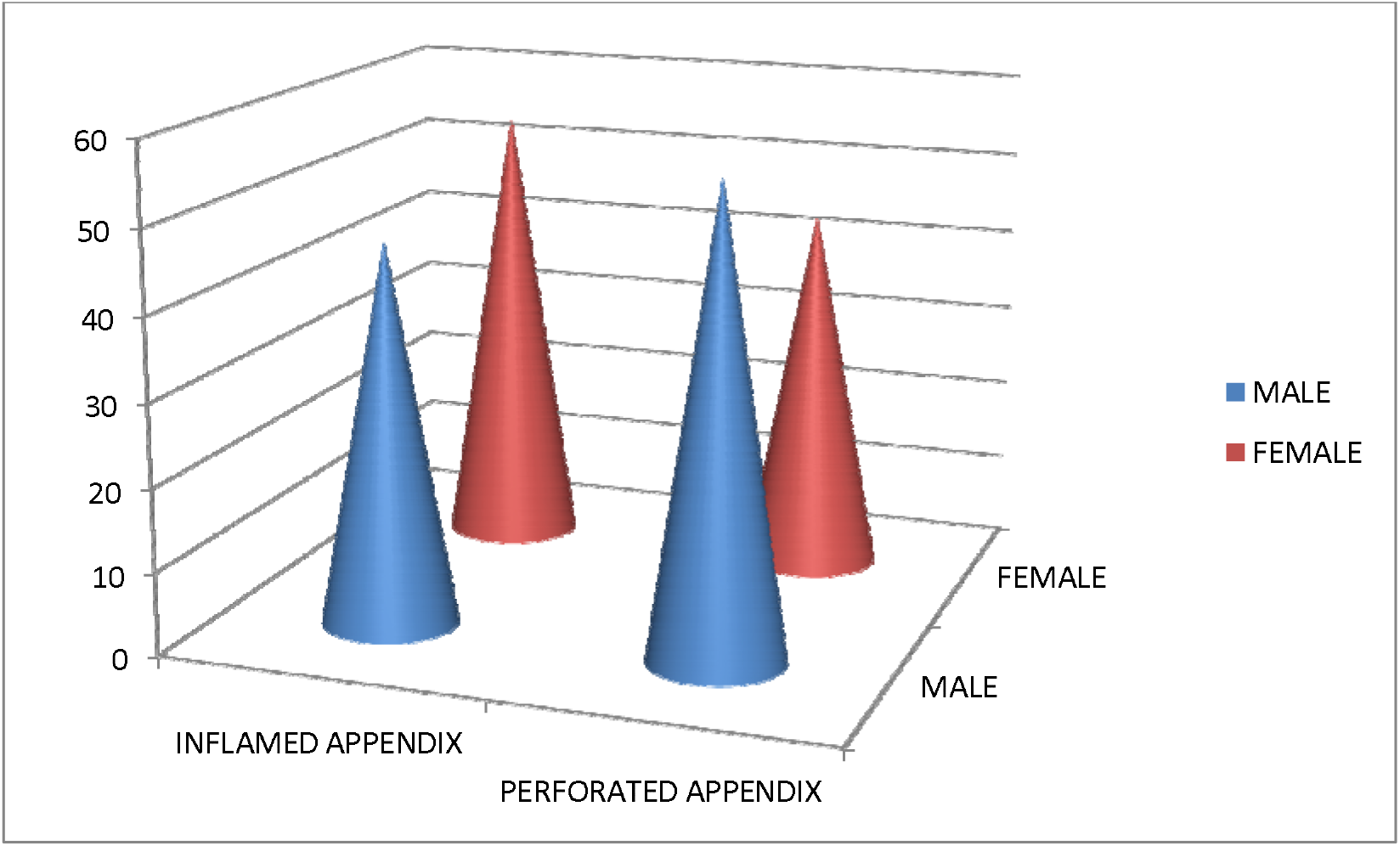
Total number of respondents with respect to their sex.

Table 1 showed that mean age was 24.13+9.73 in males and 18.7+ 6.4 in females of group A and 26.0+10.1 in males and 20.56+7.53 in females of Group B.

**Table no 1:** Mean age of respondents according to their sex

| Mean age | Inflamed appendix | Perforated appendix |
| --- | --- | --- |
| Male | 24.13+9.73 | 26.0+10.1 |
| female | 18.7+ 6.4 | 20.56+7.53 |

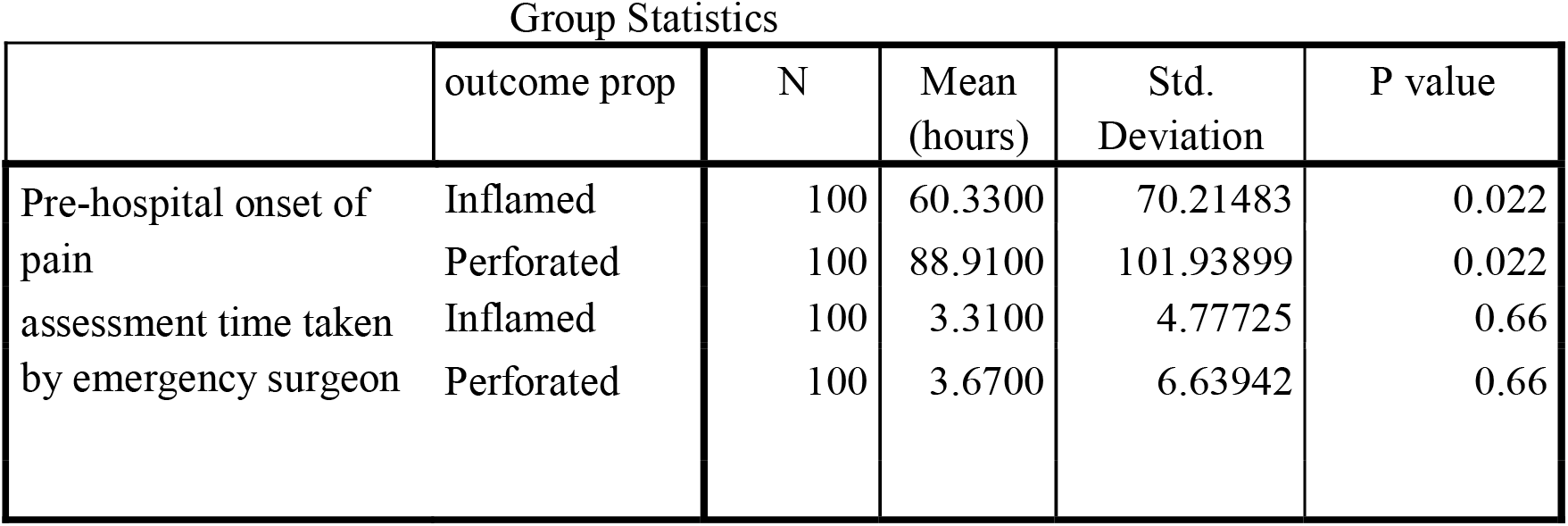

Mean duration of pain before presenting to hospital was found to be significantly longer in those with perforated appendix (p value <0.05). Time taken by surgical resident to establish a clinical diagnosis of appendicitis after patient presented in emergency department was also evaluated. No significant difference was found in In-hospital delay in diagnosis of both groups.

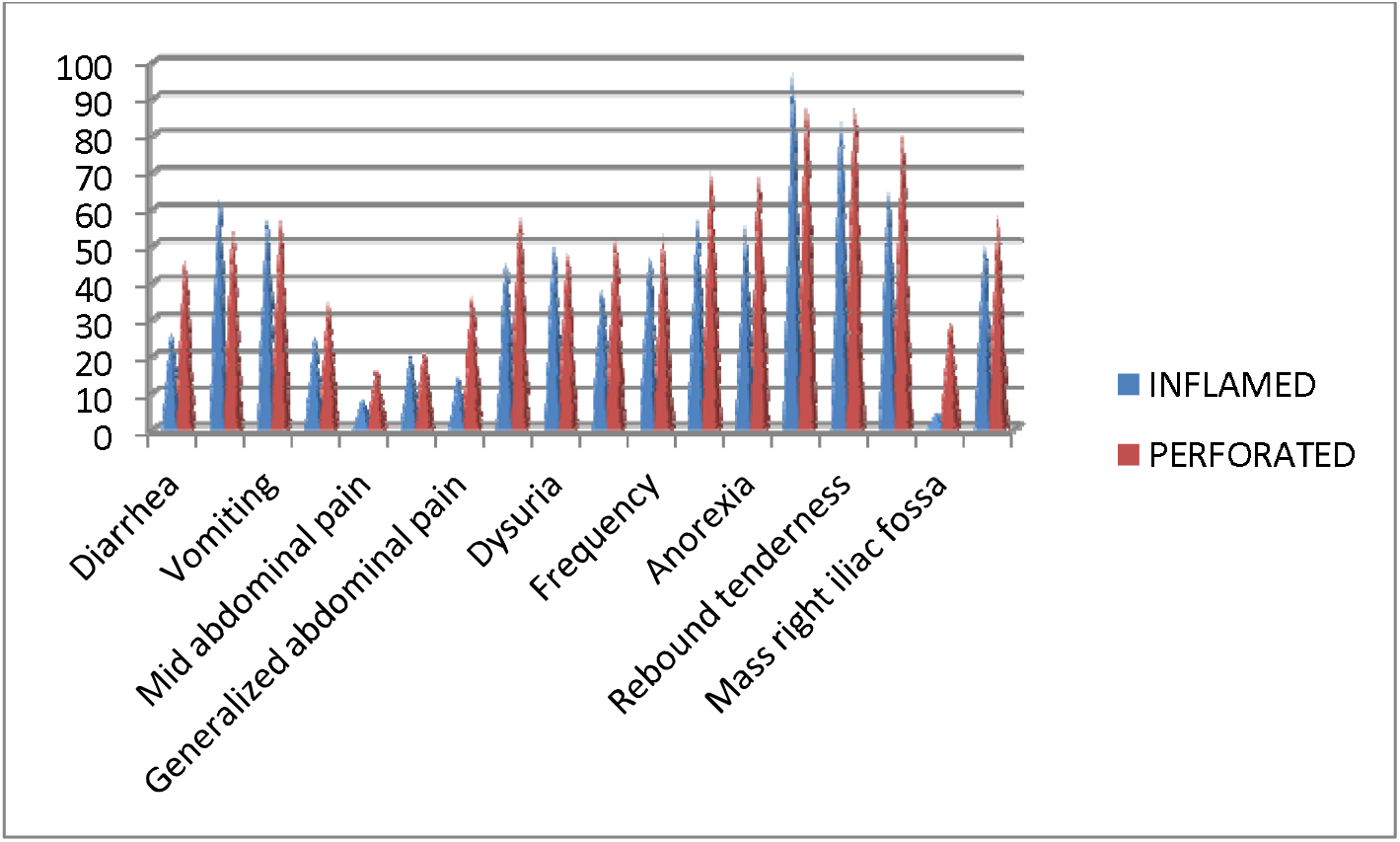

Nineteen factors in history and clinical examination were evaluated in both groups. Chi-square test showed that diarrhea, anorexia, malaise, right lumbar quadrant pain, generalized abdominal pain, tenderness and guarding in right iliac fossa on palpation, mass in right iliac fossa were significantly related to perforation. Whereas gender, nausea, fever, vomiting, pain in right iliac fossa, or dysuria were not found to be significant.

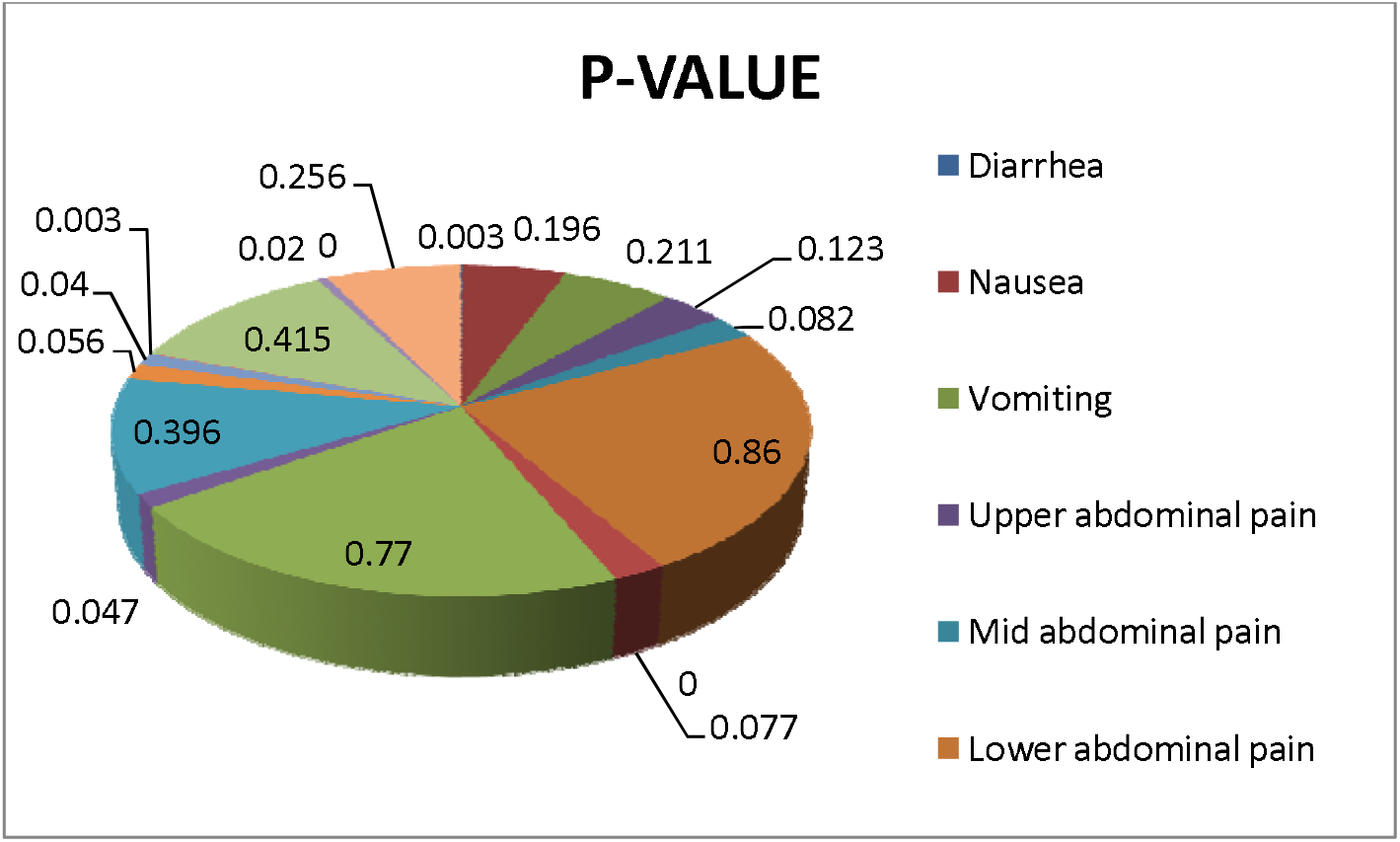

TLC was found to be significantly associated with perforation.

|  | outcome per-op | Mean | Std. Deviation | p-value |
| --- | --- | --- | --- | --- |
| total leukocyte count | inflamed | 9.8910 | 2.65654 | 0.00 |
|  | perforated | 13.0411 | 4.06697 | 0.00 |

## DISCUSSION

Acute appendicitis is the most common cause of intra-abdominal surgery in the emergency setting, the world over. The lifetime risk of suffering from this condition is approximately 8%.(10) Historically it was believed that all cases of appendicitis if untreated will lead to perforation, gangrene and abscess formation. This concept changed at the turn of the century. Nowadays the term appendicitis is used as an umbrella term covering many situations from self-limiting episodes of inflammation to frank perforation and gangrenous appendix. No specific underlying mechanism has been explained for the fulminant course of disease in the latter cases.

The current body of literature shows promising results of conservative management in cases of mild acute appendicitis(2). However, in cases where perforation has occurred, the evidence is unequivocally in favor of early surgical intervention. Therefore the importance of the ability of an emergency resident in discriminating against these two situations cannot be overestimated(11).

Over the years several scoring systems have been devised to distinguish between these two entities. Various studies show CRP, neutrophil ratio, serum bilirubin CT scan to be very useful in the early and confident diagnosis of perforated appendicitis(12,13,14). However, all these modalities are expensive and mostly unavailable in emergency setups of third world countries. Therefore, the age-old tools of history taking and bedside examination remain extremely useful in picking up cases of perforated appendicitis. In their 2010 guidelines, the American College of Emergency Physicians (ACEP) also recommends the use of clinical signs and symptoms in stratifying patients suspected of acute appendicitis (15). This helps to hierarchize patients for surgery on crammed OR lists and forethinking incision for surgery.

In this study, 19 components of history and clinical examination were investigated. Among laboratory tests, only total leucocyte count was considered that is an affordable AND EASILY AVAILABLE test at even basic health facilities.

Males have been reported present more often with perforated appendixes(16). However, in our study, no significant dominance of either gender was observed (p-value). There was no significant difference in age was also found in both age groups. This is in contrast with existing studies found (17) On history taking anorexia, nausea, generalized abdominal pain, and right lumbar quadrant pain was more significant in the perforated appendix group. On clinical examination, tenderness, guarding and rigidity were more pronounced in group b. Palpation of a phlegmonous mass in right iliac fossa was also found in 24 cases of this group (p-value). Surprisingly rebound tenderness was not able to significantly differentiate between inflamed and perforated appendix. Raised TLC and tachycardia are two important components of the classical Alvarado s score. They were raised in all cases but significantly more so in group B (p-value < 0.05). In 2016, a study by Naderan et al also found older age, diarrhea, malaise, and right lumber quadrant pain to be associated with perforated appendix(18).

Acute appendicitis is commonly believed to occur as a result of luminal obstruction. This can be due to lymphoid hyperplasia, fecalith, parasite or a malignant or benign stricture at the base. All these are believed to cause luminal obstruction leading to intraluminal fluid accumulation and resultant venous congestion. The process if left unchecked might lead to perforation usually at the tip(19,20). In this study group, A which had perforated appendix had a significant delay in hospital presentation from the onset of symptoms (p-value 0.22). the findings are supported by a few similar studies in the past as well(21). (Especially in elders and children)

In this study, the meantime from hospital presentation to open appendectomy i.e in-hospital stay time was 3.31+4.47 hours in Group A and 3.36+6.64 hours in group B (p-value 0.68). This time was consumed in assessment by the resident surgeon in emergency, baseline investigations, completion of NPO. Crammed OR lists also were responsible for some delays. This in-hospital waiting time for a few hours has been previously investigated and found to play no role in disease progression. Current recommendations state that delaying late-night presentations to the next day's list result in no adverse events. While this holds for all equivocal cases, authors believe that in all overt cases of peritonitis, early intervention might have a positive role in limiting postoperative SIRS(16).

## CONCLUSION

Despite diagnostic advancements, traditional bedside evaluation remains a relevant tool in the diagnosis of perforated appendicitis. Moreover, longer duration of pain and presence of atypical symptoms like right lumbar quadrant pain, anorexia and diarrhea should alert the treating surgeon as to the possibility of a perforated appendix.

## Data Availability

Yes, all data is with the corresponding author, it can be made available if and when required.

## Notes

### Competing Interest Statement

The authors have declared no competing interest.

### Clinical Trial

N/A

### Funding Statement

No funding

